# The Efficacy of Alpha-Lipoic Acid and B Vitamins in the Management of Diabetic Patients with Symptomatic Distal Symmetric Polyneuropathy: A Randomized Controlled Trial

**DOI:** 10.1101/2025.09.22.25336384

**Authors:** Che Nur Ain Che Abdullah, Noraini Mohamad, Nani Draman, Zainab Mat Yudin, Wan Muhamad Amir W Ahmad, Ritzzaleena Rosli Mohd Rosli, Chee Yong Chuan

**Author notes:** Corresponding author: <u>(NM);</u> (ND). These authors contributed equally to this work (Nani Draman, Zainab Mat Yudin, Wan Muhamad Amir W Ahmad, Ritzzaleena Rosli Mohd Rosli). These authors also contributed equally to this work.

## Abstract

**Background:** Distal symmetric polyneuropathy (DSPN) is a microvascular complication of diabetes that significantly impairs quality of life. However, effective treatment of symptomatic DSPN remains a challenge. This study investigates the efficacy of a fixed-dose combination of alpha lipoic acid (ALA) and B vitamins in treating type 2 diabetes mellitus(T2DM) patients with symptomatic DSPN.

**Methods:** A double-blind, randomized, placebo-controlled trial was conducted, enrolling 80 T2DM with DSPN into two groups. The intervention group received a fixed dose combination of ALA and B vitamins (*n*= 40), and the control group received a placebo (*n*=40). The efficacy was determined by using a total symptoms score (TSS), neuropathy symptom score (NSS) and neuropathy disability score (NDS) at baseline, 6 weeks, and 12 weeks. Safety profile [renal function test (RFT) and liver function test (LFT)] was measured at baseline and 12 weeks.

**Results:** A total of 80 respondents who initially participated in the study, 74 completed the 12-week treatment period; 38 in the intervention group and 36 in the placebo group. Per-protocol analyses revealed the intervention group demonstrated a significant reduction in TSS mean scores from baseline (6.92±2.76) to 6 weeks (3.51±2.57) and 12 weeks (2.45±1.92). Specifically, the mean score of TSS components in the intervention group, stabbing pain, paraesthesia, and numbness, was significantly decreased. Similarly, NSS mean score in the intervention group was significantly reduced from baseline (7.50 ± 1.16) to 6 weeks (5.29 ± 2.47) and 12 weeks (4.11 ± 2.44). NDS mean score in the intervention group also significantly reduced from 8.37 ± 1.58 at baseline to 7.08 ± 1.99 at 12 weeks. No significant changes were observed in safety parameters (RFT, LFT) after 12 weeks of treatment.

**Conclusion:** The 12-week oral treatment with a fixed-dose combination of ALA and B vitamins is effective in treating symptomatic DSPN and safe. Trial registration: ClinicalTrials.gov (NCT06568185).

## Introduction

Diabetic peripheral neuropathy (DPN) is a well-known microvascular complication of type 2 diabetes mellitus (T2DM) attributed to chronic hyperglycemia[1, 2].DPN may be focal or diffuse, with distal symmetric polyneuropathy (DSPN) being the most common diffuse type[2]. DSPN can be diagnosed by using various clinical scoring, including the neuropathy symptom score (NSS) and neuropathy disability score (NDS) [3] or Michigan neuropathy screening instrument (MNSI)[4].

Globally, the prevalence of DSPN varies widely, ranging from 30% to 40% among T2DM patients, depending on diagnostic criteria and the population studied[5, 6]. As the burden of DSPN is high, it is responsible for morbidity and disability among diabetic patients [5] and is a leading cause of disability worldwide[7]. It affects the quality of life by causing chronic pain and impaired balance, which consequently increases the risk of falls[8], a high rate of foot ulcers and amputations[9].

Despite numerous complications that may arise from DSPN, the effective treatment of symptomatic DSPN remains a challenge. First-line pharmacotherapy options for painful DSPN include several antidepressants (e.g., duloxetine, venlafaxine, amitriptyline, and other tricyclic drugs) and the gabapentinoid antiseizure medications (pregabalin, gabapentin) [10, 11]. All of them seem to be better than a placebo in alleviating pain, but they have several adverse side effects, such as nausea, somnolence, dizziness, decreased appetite, constipation, diaphoresis, and sexual dysfunction[10]. Thus, for patients who are intolerant of first-line pharmacotherapies, treatment with oral pathogenetically oriented pharmacotherapy of alpha-lipoic acid (ALA) and benfotiamine (vitamin B1) was suggested in the 2022 guidelines of the International Diabetes Federation[11].

ALA, also known as thioctic acid, is a potent antioxidant that mitigates oxidative stress, a major contributor to DSPN pathophysiology[12]. ALA may delay or reverse peripheral diabetic neuropathy through its multiple antioxidant properties[13]. Several randomized controlled trial studies have shown that ALA is effective in treating DSPN[14–17]. However, a systematic review on ALA effect in treating diabetic neuropathy reported inconsistent findings[18], with four trials demonstrating significant symptom improvement [19–22] and three showing no notable effect [23–25].

Similarly, B vitamins, including B1 (thiamine), B6 (pyridoxine), and B12 (cobalamin), are essential for nerve health by supporting myelin synthesis, nerve repair, and neurotransmitter function[26]. A systematic review concluded that B vitamin supplementation could improve many symptoms and signs of DPN [27]. However, another review article concluded that the overall efficacy of B vitamins in treating diabetic neuropathy remains uncertain and requires further study[28].

Given the inconsistent evidence regarding the individual efficacy of ALA and B vitamins in managing symptomatic DSPN, further investigation is warranted. Thus, this study aims to evaluate the efficacy of a fixed-dose combination of ALA and B vitamins in treating T2DM patients with symptomatic DSPN. Exploring this combination is important to determine whether a synergistic effect exists, with the antioxidant properties of ALA [12] may complement the neurotrophic properties provided by B vitamins that are essential in sustaining neuronal survival through multiple protective and regenerative mechanisms[29]. This combination may offer enhanced clinical benefits, contributing to optimized therapeutic strategies and improved patient outcomes in the management of DSPN.

## Methods

### Study Design

This study employed a single-center, randomized, double-blind, placebo-controlled trial design. It was conducted in accordance with the Declaration of Helsinki[30] and following the approval granted by the Human Research Ethics Committee of Universiti Sains Malaysia (USM/JEPeM/KK/23110893). This study’s protocol was registered with ClinicalTrials.gov (Identifier NCT06568185).To guarantee that the study met the suggested standards, this study was conducted and reported in accordance with the Consolidated Standards of Reporting Trials (CONSORT) statement guidelines[31].

### Population and sample

The respondents were recruited between 1^st^ June 2024 and 1^st^ December 2024 from the outpatient clinic and diabetic clinic at Universiti Sains Malaysia Specialist Hospital. Adults aged 18 years and older, with underlying T2DM who were diagnosed with DSPN using the NSS and NDS criteria were included in this study[3]. The exclusion criteria consisted of the following: participants with a documented mental impairment, peripheral vascular disease (non-palpable foot pulses, intermittent claudication), amputated foot or leg, aspartate aminotransferase or alanine aminotransferase levels more than three times the normal levels, renal impairment (chronic kidney disease stage IV and V), and individuals using medications that could potentially influence the study results, such as antidepressants, anticonvulsants, opiates, neuroleptics, antioxidants, and particularly methylcobalamin, pyridoxine and other B complex preparations. Individuals who were pregnant, lactating, or had a history of allergy to vitamin B complex preparations (i.e., Vitamin B12, B6, and B1) and ALA were also excluded. The purposive sampling method was used to select participants.

The sample size was calculated using G*Power Software version 3.1.9.7. The sample size for this study was determined based on the primary study endpoint, the change in TSS from the beginning of treatment to week 12 after treatment. Since the intended statistical analysis (repeated measures ANOVA) investigates the between-group effect and the within-group effect, the sample sizes were calculated based on two options. The largest sample size was obtained by calculating between-group effects (35 participants per group) with 80% power for detecting an effect size of 0.28 with a 95% confidence interval and Type 1 error probability of 0.05. To account for an anticipated 10% dropout rate, the final sample size was increased to 40 participants per group. Thus, the total number of participants required for this study was 80.

### Study enrolment and randomization

T2DM patients attending the outpatient and diabetes clinic were approached individually. Those who are willing to participate in this study were given a detailed explanation of the study’s significance, objectives, and procedures. The respondents were brought to the research room to be screened for their eligibility criteria, including performing NSS and NDS to determine DPN diagnosis. If all the inclusion and exclusion criteria were fulfilled, hard-copied consent was obtained from all respondents.

Then, patients were randomized into the trial in a 1:1 ratio into either the intervention group or the control group. To ensure balance between the arms, mixed block randomization was used, with independent statisticians using IBM SPSS software to assign patients to intervention and placebo groups randomly. Given the nature of the double-blind study, both respondents and clinical staff were also blinded to the study group assignments throughout the trial.

### Study medication

The intervention group received a fixed-dose combination of ALA and B vitamins (Bionerv®). Each tablet contains 300 mg of ALA, 500 mcg of methylcobalamin (vitamin B12), 8 mg of pyridoxine (vitamin B6), and 39 mg of thiamine (vitamin B1). The control group received placebo tablets consisting of croscarmellose sodium, microcrystalline cellulose, silicon dioxide, and magnesium stearate, the formulation of which was derived from the excipients used in the intervention medication. The placebo is indistinguishable in appearance, color, and texture from the active intervention tablets, ensuring effective blinding.

Respondents in the intervention and control groups need to take two tablets of medication daily after breakfast for 12 weeks. The 12-week treatment duration was selected based on prior clinical evidence, the timeline of nerve repair, safety considerations, and practical trial design constraints[32, 33].

### Study outcomes

#### Primary outcome measures

The primary outcomes of the study are the changes in NSS, NDS and TSS scores. The NSS and NDS scores are instruments used to aid in the diagnosis of DSPN. The sensitivity, specificity and diagnostic efficacy of NSS and NDS scores were checked in a study conducted by Asad et al. (2010), taking nerve conduction studies as the gold standard. It was found that NSS and NDS had 82.05% and 92.31% sensitivity and 66.67% and 47.62% specificity, respectively. The diagnostic efficacy of NSS and NDS was 77% [34].

NSS consists of four symptom components: type, location, timing, and relief by maneuver. Symptom type includes burning, numbness, or tingling scores 2 points; fatigue or cramping scores 1 point; no symptoms score 0. Symptom location, with symptoms in the feet scoring 2 points; calves 1 point; elsewhere 0 points. The third symptom component is timing: nocturnal exacerbation for 2 points; 1 point if symptoms persist day and night; 0 if only during the day. An additional point is added if symptoms wake the patient from sleep. The fourth symptom component is relief by maneuver: symptoms are reduced by either walking, scoring 2 points, or standing, 1 point, and 0 for sitting or lying down. The total score ranges from 0 to 9, categorizing DSPN as mild (3-4), moderate (5-6), or severe (7-9) [3, 6].

The NDS was derived from the neurological examination testing sensory modalities such as vibration, temperature, pin prick and Achilles tendon reflex on both feet. Vibration sense is tested with a 128 Hz tuning fork on the hallux of the big toe, scoring 0 for normal and 1 for reduced or absent on each side [35]. Normal vibration sense is defined as the patient’s ability to perceive vibration at the hallux of the big toe is similar to the reference point when the 128 Hz tuning fork is placed on the sternum or forehead. Temperature perception is evaluated with a cold tuning fork on the dorsum of the foot, following the same scoring method [36]. Pain perception is assessed using a pinprick test at the proximal end of the big toenail. Ability to distinguish sharp/blunt for each side, score 0 (sharpness present) and score 0 otherwise (sharpness absent) [35]. The Achilles tendon reflex is tested on both sides with a tendon hammer and scored 0 for normal, 1 for reflex present with reinforcement, and 2 for absent reflex [3, 6].The total NDS score ranges from 0 to 10, with severity classified as mild (3-5), moderate (6-8), or severe (9-10) [3]. The diagnosis of DSPN for this study is based on the presence of moderate symptoms (NSS > 5) with mild signs (NDS > 3) [3].

The TSS, offering a comprehensive assessment of neuropathic symptomatology, was assessed to quantify neuropathic symptom burden by summing scores for four core symptoms: stabbing pain, burning pain, paraesthesia, and numbness. Each symptom is rated based on its frequency (occasional, frequent, or continuous) and intensity (absent, slight, moderate, or severe). The combined score provides a total between 0 and 14.64, with higher TSS scores indicating greater frequency and intensity of neuropathic symptoms, reflecting a more severe symptom burden [33, 37]. The scoring method for TSS is detailed in Table 1.

**Table 1.** A scoring approach for the neuropathic symptoms included in the TSS score (stabbing pain, burning pain, paraesthesia, and numbness)

| Symptoms frequency | Symptom intensity |  |  |  |
| --- | --- | --- | --- | --- |
|  | Absent | Slight | Moderate | Severe |
| Occasional | 0 | 1.00 | 2.00 | 3.00 |
| Frequent | 0 | 1.33 | 2.33 | 3.33 |
| Almost | 0 | 1.66 | 2.66 | 3.66 |

All neurological assessments were performed by two researchers who had been trained and standardised by a neurologist, ensuring consistency in assessment techniques.

#### Secondary outcome measures

6 mL of venous blood was drawn at baseline visit to assess safety parameters, including renal function test (RFT), which measures serum creatinine, and liver function test (LFT), which assesses aspartate transaminase (AST), alanine transaminase (ALT) and alkaline phosphatase (ALP). In addition, baseline haemoglobin A1c (HbA1c) was taken.

### Follow-up visit

Follow-up visits were done at 6 and 12 weeks to reassess NSS and TSS. For NDS, LFT and RFT, it was reassessed again at 12 weeks to evaluate changes in outcomes. Other assessments include treatment compliance and documenting any adverse effects.

### Compliance and safety measures

A daily checklist form was provided to the respondents to monitor their compliance with the study medication. After taking the fixed-dose combination of ALA and B vitamins or placebo, respondents were required to mark the checklist accordingly. Compliance was defined as the consumption of at least 80% of the assigned medication throughout the study period. Non-compliant participants will be discontinued from the study and will not be replaced.

Safety measures included the monitoring of serious adverse events (SAEs) and adverse events. SAE and adverse events were monitored throughout the entire study. Participants were instructed to report any adverse effects, and this information was recorded using a standardized adverse effect reporting sheet. Any adverse events related to the study intervention were reported to the clinic and attended by a medical doctor from the research team if any treatment was needed. Participants who developed SAEs will be discontinued from the study and will not be replaced.

### Statistical Analysis

Data analysis was conducted using SPSS version 29. All randomized subjects who received at least one of the investigational products were included in the analysis. Analyses based on the intent-to-treat (ITT) principle were performed to examine baseline demographic information, medical profiles, clinical characteristics and blood parameters of respondents. The chi-square test was used to assess the distributional differences of each categorical variable (Table 2), and an independent t-test (Table 3) was applied to assess any differences between the placebo and intervention groups.

**Table 2.**
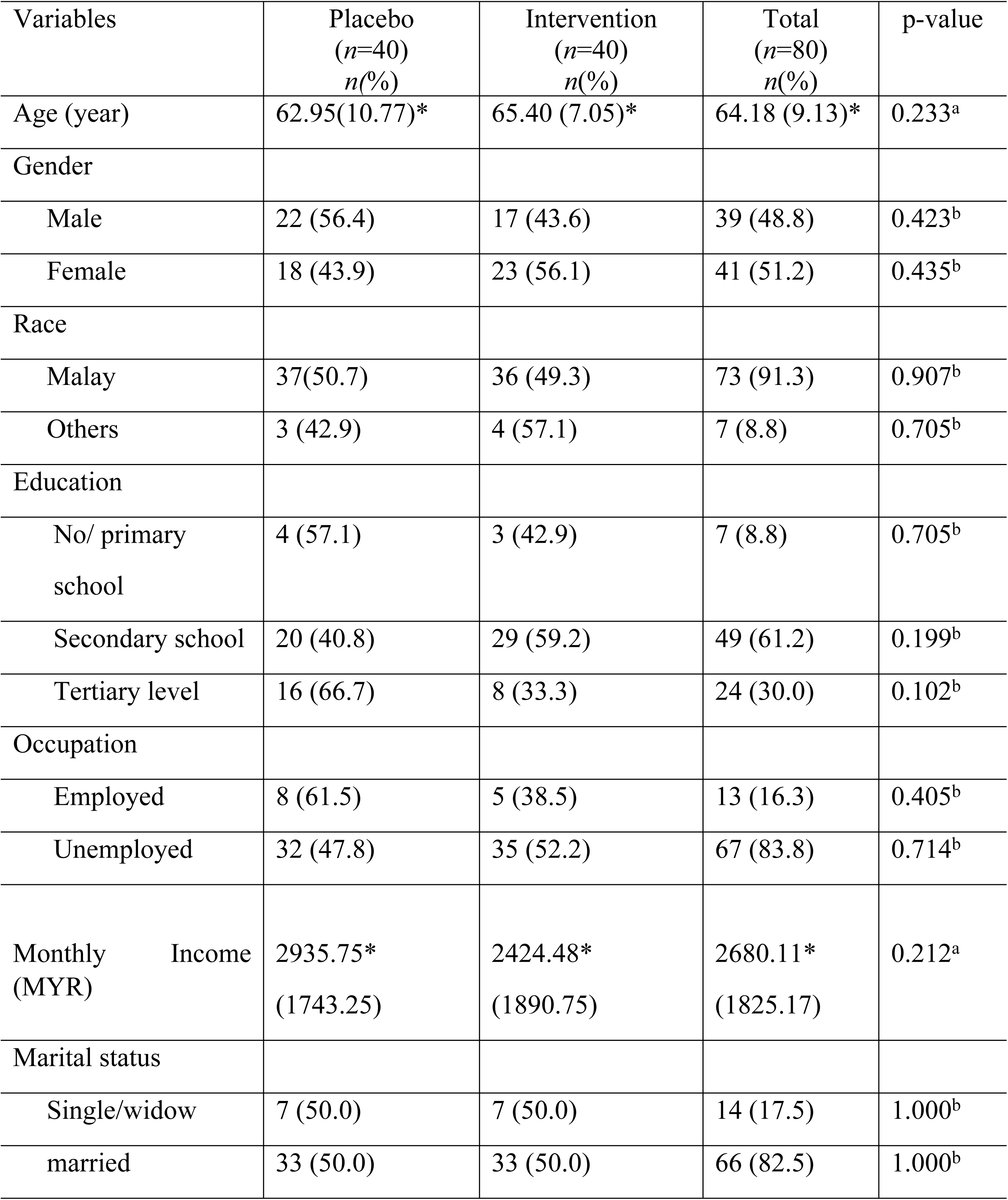
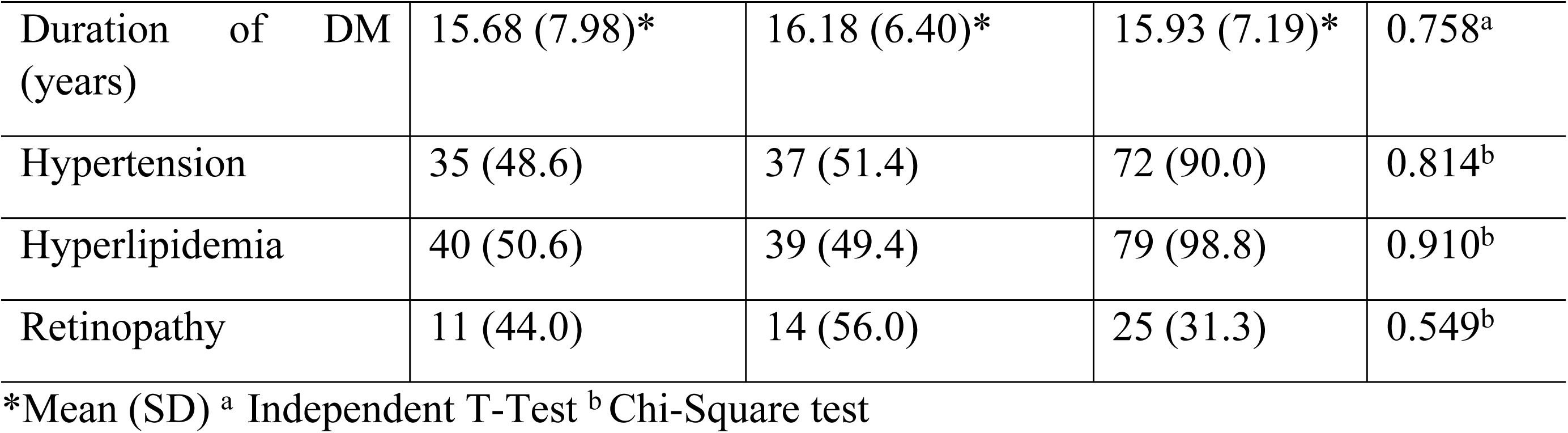
Baseline sociodemographic data and medical profile of the respondents (*n*=80)

**Table 3.**
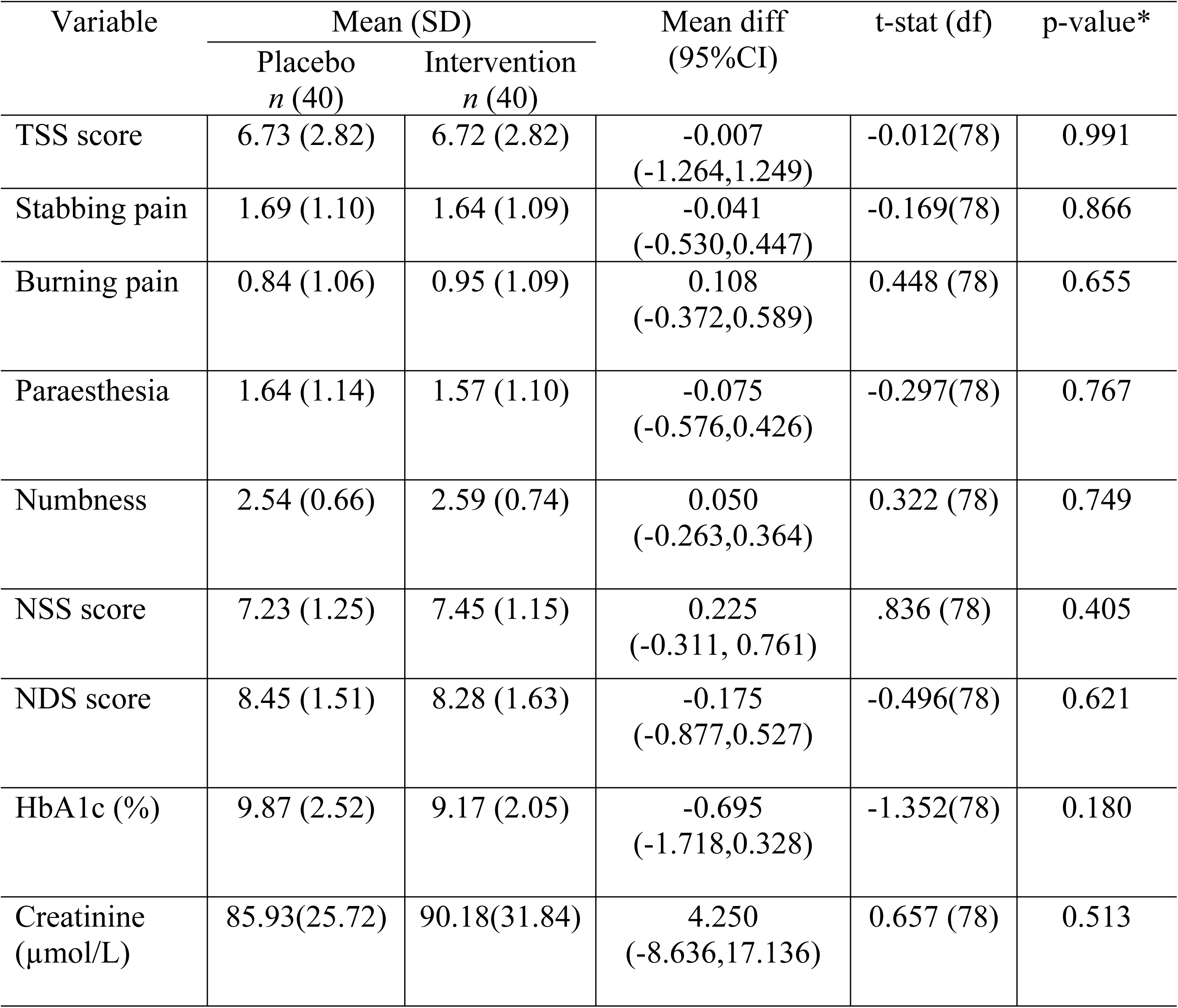
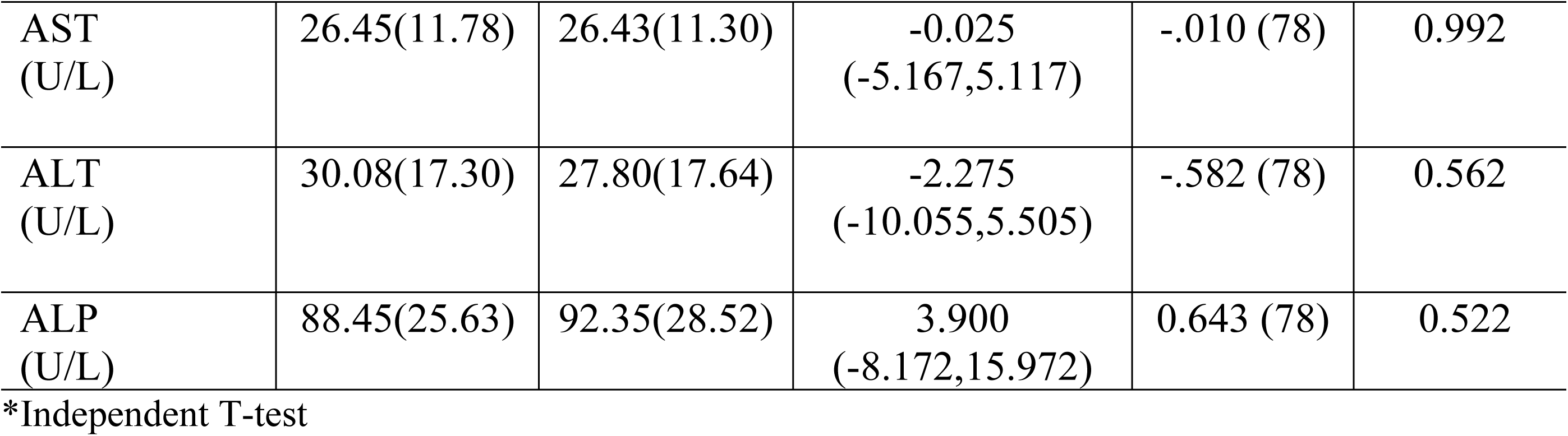
Baseline clinical characteristics and blood parameters of respondents(*n*=80)

| Variable | Mean (SD) |  | Mean diff<br>(95%CI) | t-stat (df) | p-value* |
| --- | --- | --- | --- | --- | --- |
|  | Placebo<br>n (40) | Intervention<br>n (40) |  |  |  |
| TSS score | 6.73 (2.82) | 6.72 (2.82) | -0.007<br>(-1.264,1.249) | -0.012(78) | 0.991 |
| Stabbing pain | 1.69 (1.10) | 1.64 (1.09) | -0.041<br>(-0.530,0.447) | -0.169(78) | 0.866 |
| Burning pain | 0.84 (1.06) | 0.95 (1.09) | 0.108<br>(-0.372,0.589) | 0.448 (78) | 0.655 |
| Paraesthesia | 1.64 (1.14) | 1.57 (1.10) | -0.075<br>(-0.576,0.426) | -0.297(78) | 0.767 |
| Numbness | 2.54 (0.66) | 2.59 (0.74) | 0.050<br>(-0.263,0.364) | 0.322 (78) | 0.749 |
| NSS score | 7.23 (1.25) | 7.45 (1.15) | 0.225<br>(-0.311, 0.761) | .836 (78) | 0.405 |
| NDS score | 8.45 (1.51) | 8.28 (1.63) | -0.175<br>(-0.877,0.527) | -0.496(78) | 0.621 |
| HbA1c (%) | 9.87 (2.52) | 9.17 (2.05) | -0.695<br>(-1.718,0.328) | -1.352(78) | 0.180 |
| Creatinine<br>(μmol/L) | 85.93(25.72) | 90.18(31.84) | 4.250<br>(-8.636,17.136) | 0.657 (78) | 0.513 |
| AST<br>(U/L) | 26.45(11.78) | 26.43(11.30) | -0.025<br>(-5.167,5.117) | -.010 (78) | 0.992 |
| ALT<br>(U/L) | 30.08(17.30) | 27.80(17.64) | -2.275<br>(-10.055,5.505) | -.582 (78) | 0.562 |
| ALP<br>(U/L) | 88.45(25.63) | 92.35(28.52) | 3.900<br>(-8.172,15.972) | 0.643 (78) | 0.522 |

\*Independent T-test

Additionally, per-protocol analyses were conducted to examine the robustness of the efficacy outcomes. Subjects who had major protocol deviations, such as noncompliance with the study drug, were excluded from the per-protocol analysis. A repeated measures analysis of variance (ANOVA) was conducted to assess significant changes in TSS scores, TSS components (stabbing pain, burning pain, paraesthesia, numbness), and NSS score at baseline, 6 weeks, and 12 weeks of treatment. The changes in NDS score and safety profile of RFT and LFT were measured at baseline and 12 weeks of treatment. The analysis aimed to examine: 1) within-group changes over time (time effect), and 2) between-group differences (treatment effect). All estimates were reported as estimated marginal means with adjusted 95% confidence intervals (CI). Two-tailed P-values of less than 0.05 were considered statistically significant.

### Ethical Approvals

The study protocol was reviewed and approved by the Human Research Ethics Committee, Universiti Sains Malaysia (USM/JEPeM/KK/23110893) on 31st March 2024. Hardcopy written informed consent was obtained from all participants prior to their inclusion in the study.

## Results

The subject distribution throughout the trial is shown in Fig 1. Of the 82 respondents initially screened for this study, 80 were eligible for randomization, and two did not meet the inclusion criteria due to high serum creatinine levels. Consequently, a total of 80 respondents entered the run-in phase. During the intervention period, 74 respondents completed the trial, and 6 (7.5%) discontinued participation at week six due to adverse events of medication. Specifically, two respondents (5%) from the intervention group and four (10%) respondents from the control group (Figure 1).

**Fig 1.**
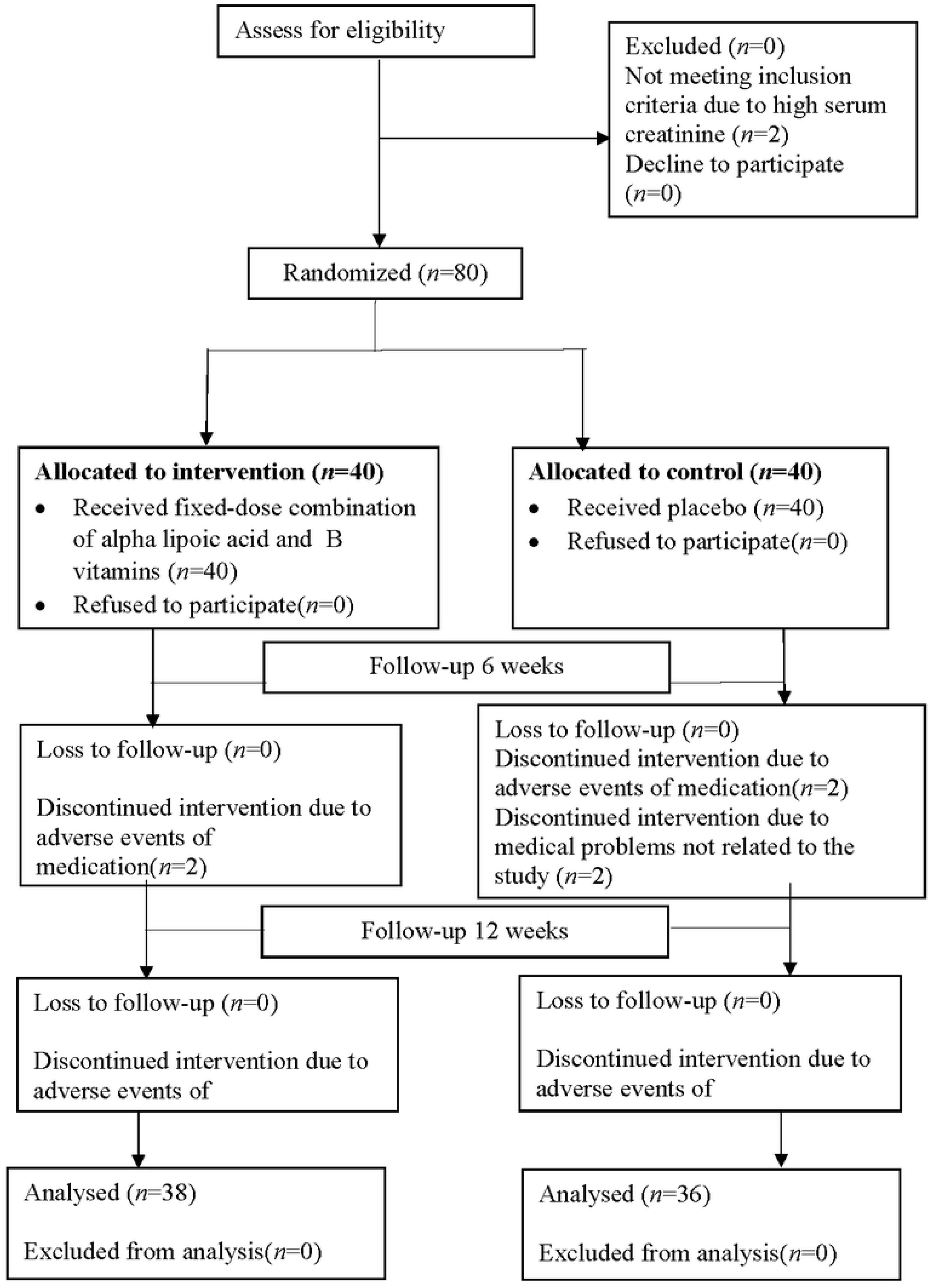
The subject dist1ibution throughout the t1ial based on the CONSORT flow.

The baseline respondents’ sociodemographic data and medical profiles are shown in Table 2. There were no significant differences between the groups in baseline sociodemographic characteristics, and the medical profiles of the respondents suggest the data were homogeneous (p>0.05).

There were also no significant differences among the groups for the baseline clinical characteristics and blood parameters of respondents, with the mean of TSS, stabbing pain, burning pain, paraesthesia, numbness, NSS, NDS, HbA1c, serum creatinine, ALT, AST and ALP score being similar in both groups with p > 0.05 (Table 3).

The final outcomes were analyzed using a per-protocol approach; i.e., only data from patients who completed 12 weeks of the randomized treatment were considered for the present analyses[33]. Therefore, the final analysis for the primary outcome was based on 74 participants, with 38 in the intervention group and 36 in the placebo group. The comparison between TSS, NSS, and NDS scores between the intervention and placebo groups at baseline, 6 weeks and 12 weeks after treatment is shown in Table 4. Based on the repeated measures ANOVA analysis, the findings can be categorized into two effects: within-group effects and between-group effects. For the within-group effect (comparing visits at baseline, 6 weeks, and 12 weeks), there was a significant change in the mean TSS score [F(df) = 89.19 (2,144), p< 0.001] and TSS components, NSS [F(df) = 39.35 (2,144); p <0.001] and NDS score [F(df) = 12.95 (2,144); p <0.001] in the intervention group.

**Table 4.**
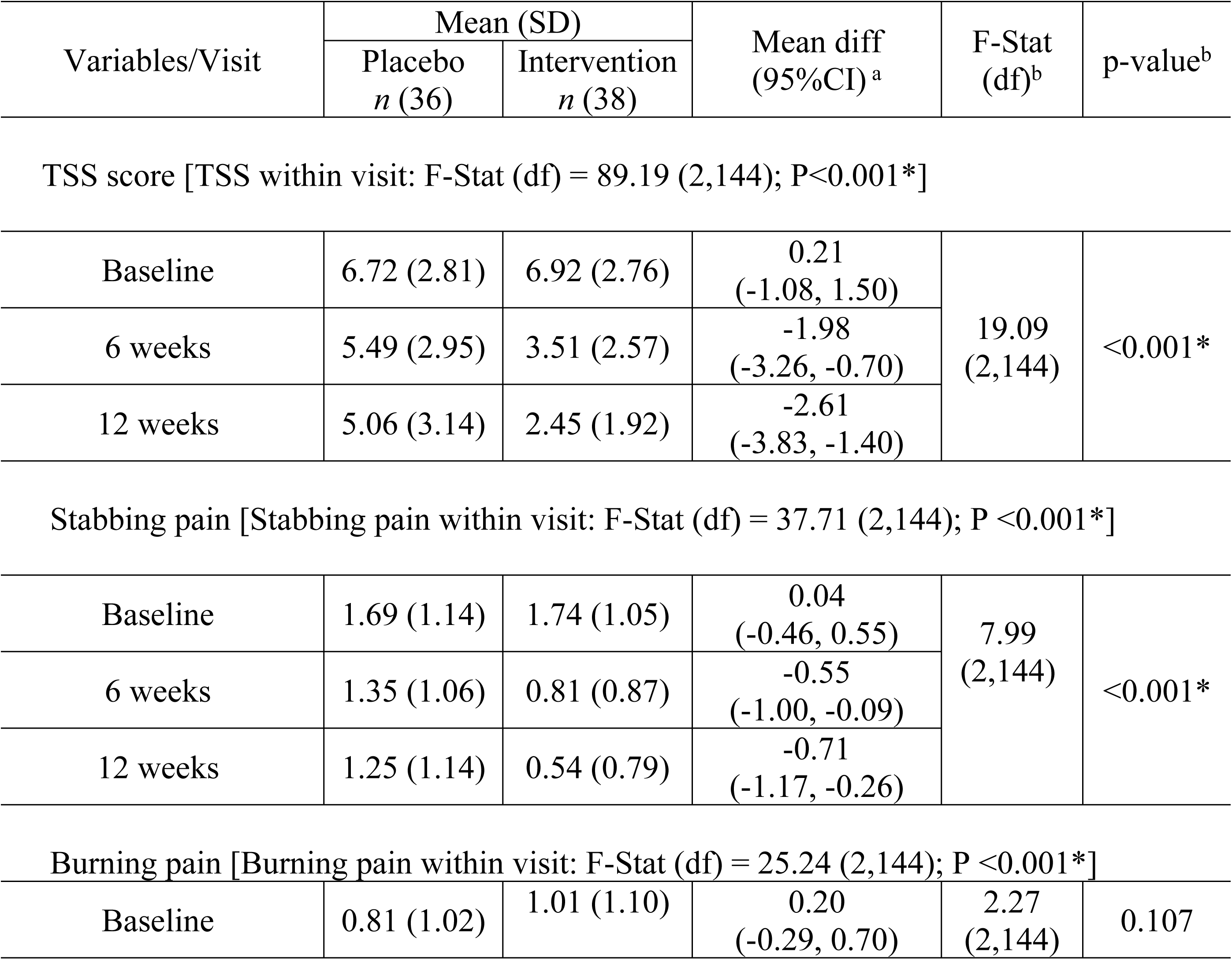
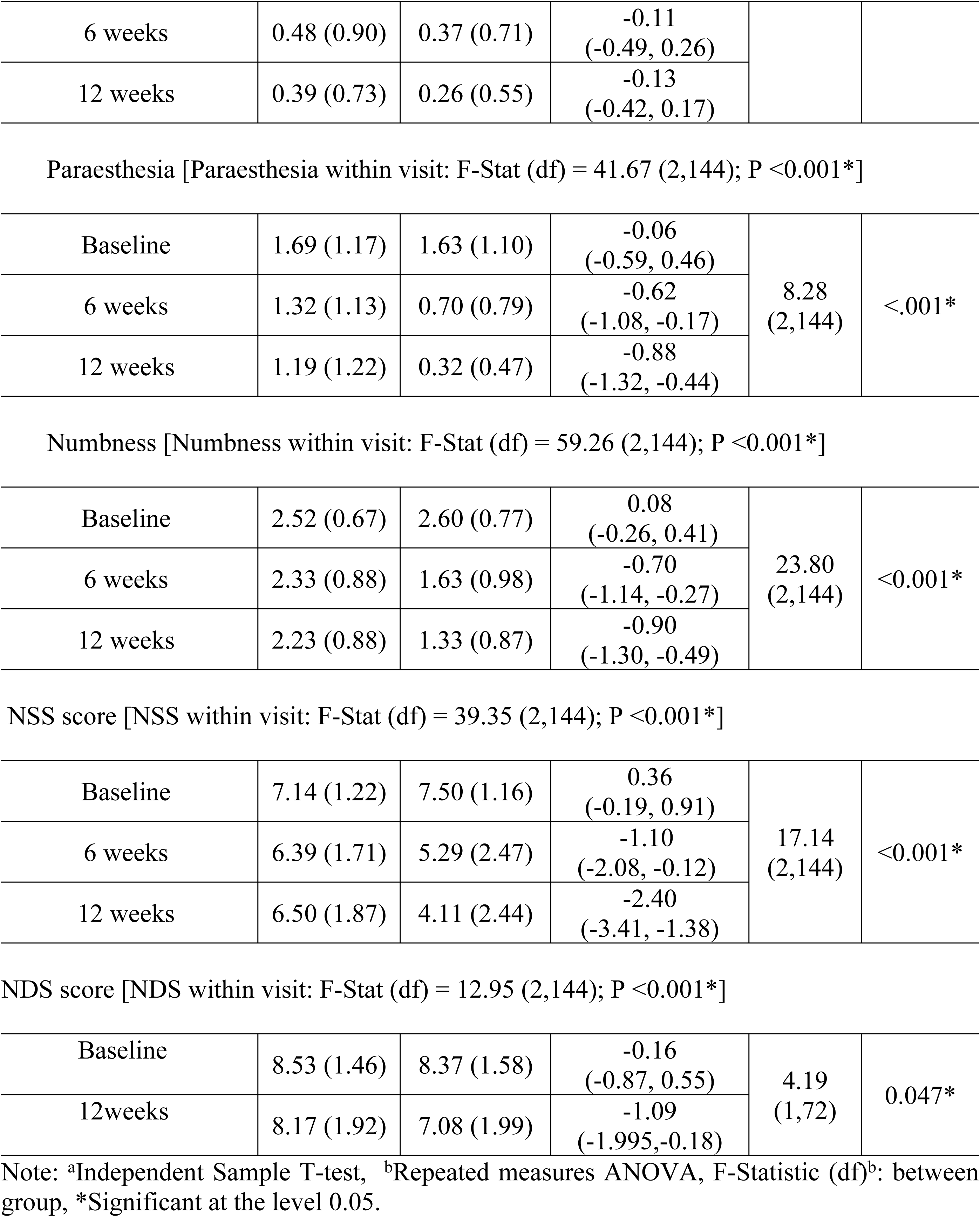
Comparison of TSS, NSS and NDS scores between intervention and placebo groups at baseline, 6 and 12 weeks after treatment (*n*=74).

| Variables/Visit | Mean (SD) |  | Mean diff<br>(95%CI)<sup>a</sup> | F-Stat<br>(df)<sup>b</sup> | p-value<sup>b</sup> |
| --- | --- | --- | --- | --- | --- |
|  | Placebo<br>*n* (36) | Intervention<br>*n* (38) |  |  |  |
| TSS score [TSS within visit: F-Stat (df) = 89.19 (2,144); P<0.001*] |  |  |  |  |  |
| Baseline | 6.72 (2.81) | 6.92 (2.76) | 0.21<br>(-1.08, 1.50) | 19.09<br>(2,144) | <0.001* |
| 6 weeks | 5.49 (2.95) | 3.51 (2.57) | -1.98<br>(-3.26, -0.70) |  |  |
| 12 weeks | 5.06 (3.14) | 2.45 (1.92) | -2.61<br>(-3.83, -1.40) |  |  |
| Stabbing pain [Stabbing pain within visit: F-Stat (df) = 37.71 (2,144); P <0.001*] |  |  |  |  |  |
| Baseline | 1.69 (1.14) | 1.74 (1.05) | 0.04<br>(-0.46, 0.55) | 7.99<br>(2,144) | <0.001* |
| 6 weeks | 1.35 (1.06) | 0.81 (0.87) | -0.55<br>(-1.00, -0.09) |  |  |
| 12 weeks | 1.25 (1.14) | 0.54 (0.79) | -0.71<br>(-1.17, -0.26) |  |  |
| Burning pain [Burning pain within visit: F-Stat (df) = 25.24 (2,144); P <0.001*] |  |  |  |  |  |
| Baseline | 0.81 (1.02) | 1.01 (1.10) | 0.20<br>(-0.29, 0.70) | 2.27<br>(2,144) | 0.107 |
| 6 weeks | 0.48 (0.90) | 0.37 (0.71) | -0.11<br>(-0.49, 0.26) |  |  |
| 12 weeks | 0.39 (0.73) | 0.26 (0.55) | -0.13<br>(-0.42, 0.17) |  |  |
| Paraesthesia [Paraesthesia within visit: F-Stat (df) = 41.67 (2,144); P <0.001\*] |  |  |  |  |  |
| Baseline | 1.69 (1.17) | 1.63 (1.10) | -0.06<br>(-0.59, 0.46) | 8.28<br>(2,144) | <.001* |
| 6 weeks | 1.32 (1.13) | 0.70 (0.79) | -0.62<br>(-1.08, -0.17) |  |  |
| 12 weeks | 1.19 (1.22) | 0.32 (0.47) | -0.88<br>(-1.32, -0.44) |  |  |
| Numbness [Numbness within visit: F-Stat (df) = 59.26 (2,144); P <0.001\*] |  |  |  |  |  |
| Baseline | 2.52 (0.67) | 2.60 (0.77) | 0.08<br>(-0.26, 0.41) | 23.80<br>(2,144) | <0.001* |
| 6 weeks | 2.33 (0.88) | 1.63 (0.98) | -0.70<br>(-1.14, -0.27) |  |  |
| 12 weeks | 2.23 (0.88) | 1.33 (0.87) | -0.90<br>(-1.30, -0.49) |  |  |
| NSS score [NSS within visit: F-Stat (df) = 39.35 (2,144); P <0.001\*] |  |  |  |  |  |
| Baseline | 7.14 (1.22) | 7.50 (1.16) | 0.36<br>(-0.19, 0.91) | 17.14<br>(2,144) | <0.001* |
| 6 weeks | 6.39 (1.71) | 5.29 (2.47) | -1.10<br>(-2.08, -0.12) |  |  |
| 12 weeks | 6.50 (1.87) | 4.11 (2.44) | -2.40<br>(-3.41, -1.38) |  |  |
| NDS score [NDS within visit: F-Stat (df) = 12.95 (2,144); P <0.001\*] |  |  |  |  |  |
| Baseline | 8.53 (1.46) | 8.37 (1.58) | -0.16<br>(-0.87, 0.55) | 4.19<br>(1,72) | 0.047* |
| 12weeks | 8.17 (1.92) | 7.08 (1.99) | -1.09<br>(-1.995, -0.18) |  |  |

Note: <sup>a</sup>Independent Sample T-test, <sup>b</sup>Repeated measures ANOVA, F-Statistic (df)<sup>b</sup>: between group, \*Significant at the level 0.05.

For the between-group effect (comparing visits at baseline, 6 weeks, and 12 weeks between the placebo and intervention groups), there was a significant reduction in the mean score of TSS, stabbing pain, paraesthesia, numbness, NSS, and NDS in the intervention group compared to the placebo group over the 12-week treatment period. In the intervention group, the TSS means score significantly reduced from baseline (6.92 ± 2.76) to 6 weeks (3.51 ± 2.57) and 12 weeks (2.45 ± 1.92), compared to the placebo group (p<0.001) (Fig 2). Specifically, the mean score of TSS components in the intervention group showed significant reductions; 1) stabbing pain significantly reduced from baseline (1.74 ± 1.05) to 6 weeks (0.81 ± 0.87) and 12 weeks(0.54 ± 0.79) compared to the placebo group (p < 0.001), 2) paraesthesia significantly decreased from baseline (1.63 ± 1.10) to 6 weeks (0.70 ± 0.79) and 12 weeks (0.32 ± 0.47) compared to the placebo group (p < 0.001) and, 3) numbness significantly reduced from baseline (2.60± 0.77) to 6 weeks (1.63 ± 0.98) and 12 weeks (1.33 ± 0.87) compared to the placebo group (p < 0.001). The mean changes of TSS components for burning pain were decreased, although not significant, from baseline (1.01 ± 1.10) to 6 weeks (0.37 ± 0.71) and 12 weeks (0.26 ± 0.55) compared to the placebo group (p= 0.107). Meanwhile, the NSS mean scores in the intervention group reduced significantly over the 12-week period, at baseline (7.50 ± 1.16), 6 weeks (5.29 ± 2.47), and 12 weeks (4.11 ± 2.44), when compared to the placebo group (p<0.001) (Fig 3). The mean NDS score in the intervention group also reduced significantly from 8.37 ± 1.58 at baseline to 7.08 ± 1.99 at 12 weeks (p=0.047) (Fig 4).

**Fig 2.**
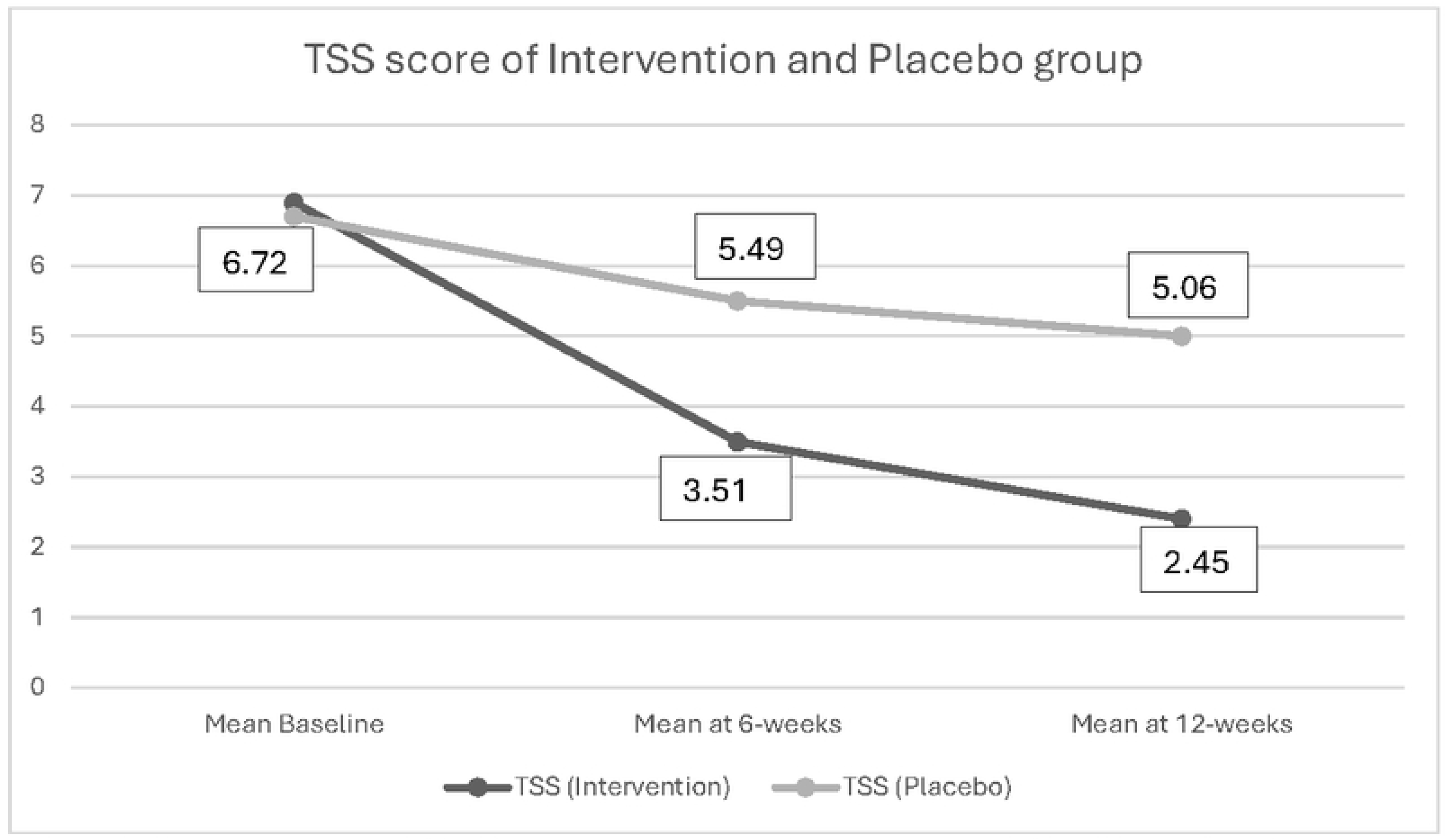
TSS means score of the intervention a11d placebo group after 12 weeks of treatment.

**Fig 3.**
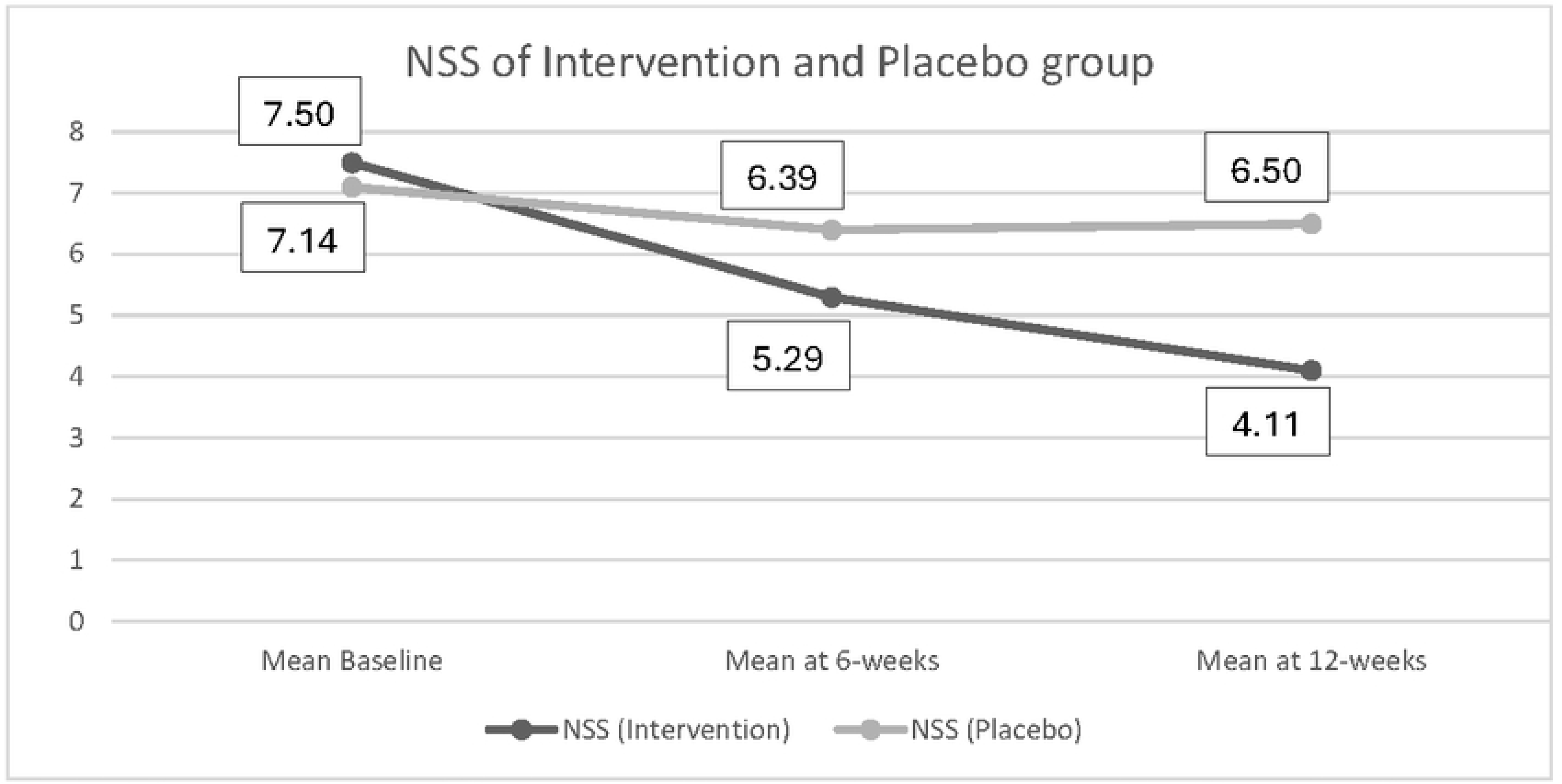
NSS means score of the intervention and placebo group after 12 weeks of treatment.

**Fig 4.**
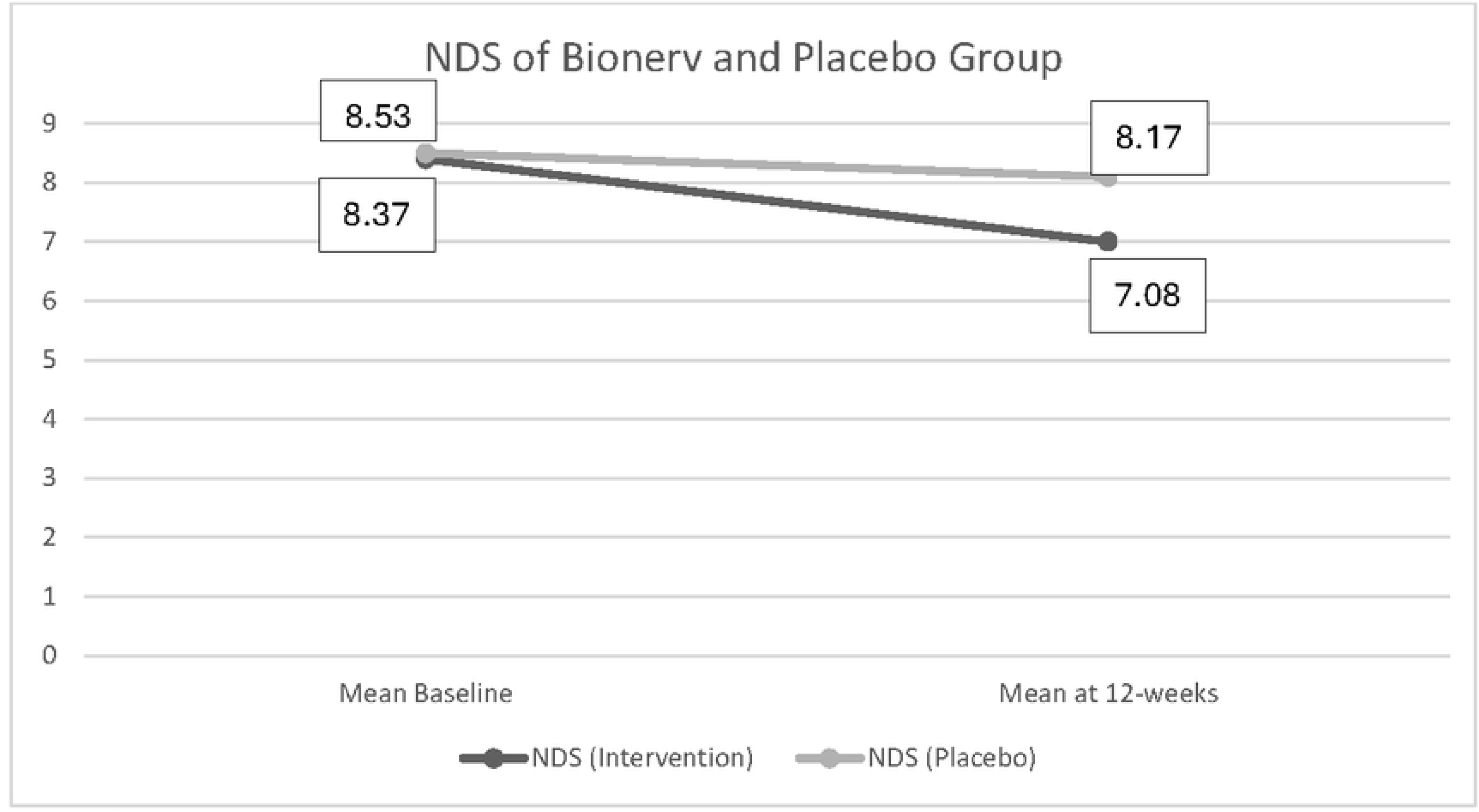
NDS means score of the intervention and placebo group after 12 weeks of treatment.

Table 5 presents safety parameters outcomes over 12 weeks. Renal and hepatic safety parameters showed no significant differences between and within groups over the study period. Serum creatinine levels remained stable in the intervention and placebo groups, with no significant changes with p=0.630. Similarly, liver enzymes (AST, ALT and ALP) showed no significant changes after 12 weeks of treatment (p> 0.05), supporting a favourable safety profile for the intervention.

**Table 5:**
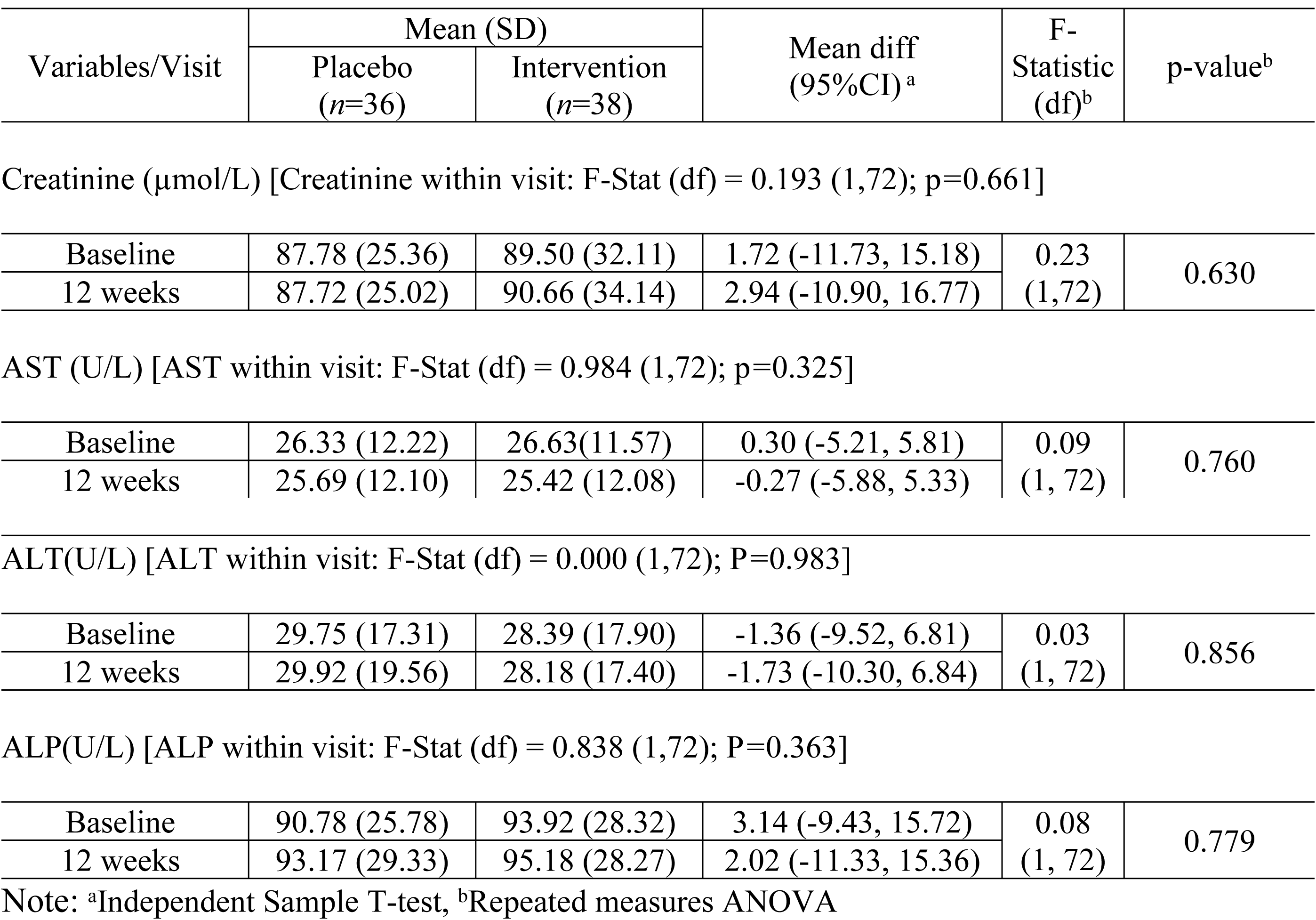
Comparison of safety parameters of intervention and placebo groups at baseline and 12 weeks after treatment (*n*=74)

| Variables/Visit | Mean (SD) |  | Mean diff<br>(95%CI)<sup>a</sup> | F-Statistic<br>(df)<sup>b</sup> | p-value<sup>b</sup> |
| --- | --- | --- | --- | --- | --- |
| | Placebo<br>($n=36$) | Intervention<br>($n=38$) | | | |
| Creatinine ( $\mu\text{mol/L}$ ) [Creatinine within visit: F-Stat (df) = 0.193 (1,72); $p=0.661$ ] |  |  |  |  |  |
| Baseline | 87.78 (25.36) | 89.50 (32.11) | 1.72 (-11.73, 15.18) | 0.23<br>(1, 72) | 0.630 |
| 12 weeks | 87.72 (25.02) | 90.66 (34.14) | 2.94 (-10.90, 16.77) |  |  |
| AST (U/L) [AST within visit: F-Stat (df) = 0.984 (1,72); $p=0.325$ ] |  |  |  |  |  |
| Baseline | 26.33 (12.22) | 26.63 (11.57) | 0.30 (-5.21, 5.81) | 0.09<br>(1, 72) | 0.760 |
| 12 weeks | 25.69 (12.10) | 25.42 (12.08) | -0.27 (-5.88, 5.33) |  |  |
| ALT(U/L) [ALT within visit: F-Stat (df) = 0.000 (1,72); $P=0.983$ ] |  |  |  |  |  |
| Baseline | 29.75 (17.31) | 28.39 (17.90) | -1.36 (-9.52, 6.81) | 0.03<br>(1, 72) | 0.856 |
| 12 weeks | 29.92 (19.56) | 28.18 (17.40) | -1.73 (-10.30, 6.84) |  |  |
| ALP(U/L) [ALP within visit: F-Stat (df) = 0.838 (1,72); $P=0.363$ ] |  |  |  |  |  |
| Baseline | 90.78 (25.78) | 93.92 (28.32) | 3.14 (-9.43, 15.72) | 0.08<br>(1, 72) | 0.779 |
| 12 weeks | 93.17 (29.33) | 95.18 (28.27) | 2.02 (-11.33, 15.36) |  |  |

Note: <sup>a</sup>Independent Sample T-test, <sup>b</sup>Repeated measures ANOVA

### Adverse event

Among the 80 study participants, 16 (20%) experienced adverse events at the six-week follow-up: 11 (27.5%) participants from the intervention group and 5 (12.5%) from the placebo group. The most frequently reported adverse event in the intervention group was nausea (15%), followed by itchiness (5%), dizziness (5%), and light-headedness (5%). The adverse event profile in the placebo group was similar, with nausea reported in 5% of participants, itchiness in 2.5%, dizziness in 2.5%, and light-headedness in 2.5%. At the 12-week follow-up, the incidence of adverse events declined, with only two participants reporting mild but tolerable nausea: one (2.5%) from the intervention group and one (2.5%) from the placebo group.

Two participants (5%) in the intervention group discontinued the study treatment due to nausea and itchiness. In the placebo group, four participants (10%) withdrew from the study; two of them discontinued due to unrelated medical conditions (one reported femur pain post-plating, and another experienced an acute exacerbation of bronchial asthma). All adverse events were assessed by the investigator and determined to have an “unlikely” causal relationship with the study medication. The remaining two participants in the placebo group discontinued due to nausea. No serious adverse events were reported in this study.

## Discussion

Symptomatic DSPN significantly impacts the quality of life for patients with T2DM[11]. Hence, effective treatment is needed to alleviate this condition. Existing individual clinical trials on ALA[17] or B vitamins preparations[27] in treating DSPN have been shown to improve the symptoms but lack robust evidence supporting their combined efficacy. This study bridges that gap by evaluating a fixed-dose combination of these therapies, hypothesizing their synergistic effects on symptom reduction in DSPN patients.

In this study, the intervention group that received a fixed-dose combination of ALA and B vitamins (B12, B6, and B1) (Bionerv®) demonstrated significant improvements across all key neuropathy-related outcomes, including TSS score, TSS score components [stabbing pain, paraesthesia, numbness, NSS score, and NDS score over 12 weeks of treatment when compared to the placebo group. A previous Korean study investigating the efficacy of oral ALA (600mg/day) in patients with painful DPN (*n*=52) found the TSS mean score significantly improved from baseline (5.15±3.57) to 12 weeks (3.52±3.39) of treatment. This study also reported that there was a significant decrease in mean sub-scores of stabbing pain(baseline 1.62 ± 1.26 to 12-weeks 1.04 ± 0.93), paraesthesia (baseline 1.27 ± 1.29 to 12-weeks 0.74 ± 1.04), and numbness(baseline 1.03 ± 1.16 to 12-weeks 0.65 ± 0.97)[33]. A study conducted in Mexico among DPN patients assessing the efficacy of ALA (600mg/day) over 16 weeks (*n*=45) found the TSS mean score decreased from baseline (8.9 ± 1.8) to 4 weeks (3.46 ± 2.0) of treatment and TSS mean score improved from 3.7 ± 1.9 to 2.5 ± 2.5 at 16 weeks of treatment [38]. Another study conducted in Pakistan among diabetic patients with TSS ≥ 4.0 points (*n*=110) receiving ALA 600mg/day reported the mean change in TSS was 2.38±1.99 as compared to the control group, which was 0.53±1.32[16].

Previous clinical trials have provided supportive evidence of the efficacy of ALA in the management of DPN. An early study, the SYDNEY 2 trial conducted in 2006, evaluated the effects of ALA (600mg/day) among diabetes patients with symptomatic DSPN (*n*=45) for a duration of 5 weeks. The study demonstrated a reduction in the TSS means score from baseline (9.44 ±1.86) to 5 weeks (4.85 ± 3.03) when compared to the placebo group. In this study, they also found that all components of the TSS mean score of stabbing pain, burning pain, paraesthesia, and numbness were significantly reduced after 5 weeks of ALA treatment (p<0.05) [14]. Subsequently, the NATHAN 1 trial conducted in 2011 to evaluate the long-term efficacy and safety of ALA(600mg/day) over a 4-year period in patients with mild to moderate diabetic distal symmetric sensorimotor polyneuropathy (*n*=460) concluded that long-term ALA treatment resulted in a clinically meaningful improvement and prevention of progression of neuropathic impairments and was well tolerated[15].

In addition to the improvements observed in the TSS scores, our study also demonstrated significant reductions in both NSS and NDS scores among patients receiving the fixed-dose ALA and B vitamins. These findings are consistent with previous studies. For example, a study in Egypt to investigate the efficacy of oral ALA 600mg twice daily over 6 months among DPN patients (*n*=200) reported significantly greater improvements in both NSS and NDS scores in the ALA-treated patients[22]. Another study conducted in Greece to evaluate the effect of ALA (600mg/day) among painful diabetic neuropathy patients (*n*=72) found the mean of NSS reduced from baseline 7.9 (range 4–10) to 5.3 (range 2–10) at day 40 of treatment. An earlier study in Germany to investigate the efficacy of ALA (600mg/day) for 3 weeks among symptomatic diabetic polyneuropathy patients (*n*=24) found the NDS decreased significantly among patients who received ALA[39].

The pathogenesis and natural history of DSPN are influenced by multiple interrelated factors[40]. Hyperglycemia and dyslipidemia are significant contributors to the onset of DSPN. Chronic hyperglycemia leads to substrate overload, resulting in the accumulation of toxic metabolites and mitochondrial malfunction, which exacerbate metabolic and oxidative stress and ultimately lead to axonal degeneration[41]. Excess glucose induces hyperactivity in the polyol and hexosamine pathways, elevating the levels of reactive oxygen species (ROS) and promoting inflammation, both of which contribute to mitochondrial damage and neuronal injury[42].

A systematic review and meta-analysis on the effect of ALA on oxidative stress parameters found that ALA supplementation may reduce lipid peroxidation and enhance antioxidant defence, thereby contributing to a reduction of ROS[43]. Therefore, ALA has been proposed as an effective intervention for treating DSPN[44]. It was suggested that for patients who cannot tolerate first-line pharmacotherapies for painful DSPN due to its side effects, and are interested in a nutritional supplement approach, ALA 600mg daily is recommended as a potent antioxidant that may alleviate oxidative stress, improve the underlying pathophysiology of neuropathy, and mitigate neuropathic pain[10].

Likewise, neurotropic B vitamins, specifically B1, B6, and B12, consistently protect nerves from detrimental environmental factors; for example, vitamin B1 functions as a site-specific antioxidant, vitamin B6 regulates nerve metabolism, and vitamin B12 preserves myelin sheaths. The presence of adequate levels of vitamins B1, B6, and B12 facilitates crucial nerve regeneration by promoting the formation of new cellular structures. Consequently, the deficiency of these vitamins can accelerate irreversible nerve deterioration and discomfort, ultimately resulting in peripheral neuropathy[29]. Study have found that, deficiency of vitamin B6 and vitamin B1 was prevalent in T2DM[45]. In addition, metformin usage interferes with calcium-dependent membrane action responsible for vitamin B12 intrinsic factor absorption in the terminal ileum. Prevalence of vitamin B12 deficiency after metformin use has been reported to be 22.2%. Concurrent supplementation of B vitamins may potentially protect against the deficiency[46].

In regard to the safety outcomes, our study found no significant differences between the intervention and placebo groups in the renal and hepatic safety parameters, with serum creatinine, ALT, AST and ALP remaining stable and unchanged. Similar findings were supported by a study done by Didangelos et al. after 12 months of treatment with ALA and vitamin B12, Elbadawy et al. after 3 months of ALA, and a long-term ALA supplementation of 600 mg/day for over 4 years conducted by Ziegler et al.([15, 17, 47]. This indicates a favourable safety profile for the ALA and B vitamins combination and was supported by the findings of previous studies [17, 47].

In this study, side effects were more frequently reported in the intervention group, with nausea being the most common (15%) at six weeks. This aligns with findings from studies on ALA, where nausea, vomiting, and vertigo were reported as common side effects[14]. The placebo group reported fewer side effects, consistent with expectations for inactive control. The mild adverse profile supports the use of ALA and B vitamins in long-term management, provided that patients are adequately counselled on potential side effects.

Despite the findings, this study has several limitations. First, according to the American Diabetes Association (ADA) statement in 2010, DSPN diagnosis required both abnormal nerve conduction studies (NCS) and the presence of symptoms or signs of neuropathy [48]. However, in the 2017 ADA statement, the diagnosis of DSPN is primarily clinical, based on typical symptoms and signs of symmetrical distal sensory loss, with electrophysiological testing reserved for atypical, uncertain cases or when another underlying cause is suspected[2, 49]. In the present study, NCS was not performed, and diagnosis of DSPN was established using validated clinical tools, namely the NSS and NDS scores, which have been widely applied to establish the diagnosis of DSPN in previous studies conducted in Brazil, Germany, the United Kingdom, and Malaysia [6, 50–52]. Future research should incorporate NCS results to strengthen diagnostic accuracy and to serve as an objective primary outcome to measure treatment efficacy. Second, baseline serum B vitamin levels were not assessed in this study. Future research should incorporate these measurements, as deficiencies in vitamins B1, B6, and B12 are recognized contributors to the pathogenesis and progression of DSPN[29]. Third, this study is a single-centre study, which may restrict generalizability, and the relatively short follow-up period, which limits understanding of long-term efficacy and safety. Moreover, the predominantly Malay cohort limits applicability to other ethnicities. Future studies should consider multi-centre designs, larger sample sizes, and longer follow-up durations to confirm the findings and assess the sustainability of benefits.

## Conclusions

This study highlights the significant benefits of a fixed-dose combination of ALA and B vitamins in managing T2DM patients with symptomatic DSPN. Over 12 weeks of treatment, the intervention group demonstrated superior improvements in symptom relief and overall neuropathy scores of TSS, NSS, and functional aspects of NDS when compared to the placebo group, with manageable side effects. There were also no significant changes in renal or hepatic parameters throughout the intervention, indicating a favorable safety profile. These findings suggest that this combination therapy effectively addresses both the symptomatic and functional aspects of DSPN, likely due to its dual mechanisms of antioxidant protection and neuroprotection. Nevertheless, while the results are promising, further research involving multi-center trials, longer follow-up periods, and diverse populations is necessary to confirm these benefits and establish the therapy as a standard component of DSPN management. In conclusion, oral treatment of the fixed-dose combination of ALA and B vitamins over 12 weeks is effective and safe in managing T2DM with symptomatic DSPN.

## Supporting information

**S1 Fig. The subject distribution throughout the trial based on the CONSORT flow diagram**

**S2 Fig. TSS means score of the intervention and placebo group after 12 weeks of treatment**

**S3 Fig. NSS means score of the intervention and placebo group after 12 weeks of treatment**

**S4. Fig. NDS means score of the intervention and placebo group after 12 weeks of treatment**

**S1 File. Consort 2010 checklist. Checklist according to the CONSORT criteria. (PDF)**

**S2 File. Clinical trial protocol. Ethical Submission February 19, 2024. (PDF)**

## Data Availability

All relevant data are within the manuscript and its Supporting Information files

## Acknowledgements

The researchers extend their sincere gratitude to all respondents and contributors who facilitated the successful completion of this study. Special appreciation is given to Dr. Anna Misyail Abdul Rashid, neurologist from Universiti Putra Malaysia, for her invaluable assistance in the preparation of the research proposal.

## Author Contributions

**Conceptualization:** Noraini Mohamad, Nani Draman, Zainab Mat Yudin

**Data curation:** Che Nur Ain Che Abdullah, Noraini Mohamad

**Formal analysis:** Che Nur Ain Che Abdullah, Noraini Mohamad, Muhamad Amir W Ahmad

**Funding acquisition:** Noraini Mohamad

**Investigation:** Che Nur Ain Che Abdullah, Noraini Mohamad

**Methodology:** Noraini Mohamad, Nani Draman, Muhamad Amir W Ahmad, Ritzzaleena Rosli Mohd, Chee Yong Chuan

**Supervision:** Noraini Mohamad, Nani Draman, Zainab Mat Yudin, Ritzzaleena Rosli Mohd Rosli, Chee Yong Chuan

**Writing – original draft:** Che Nur Ain Che Abdullah

**Writing – review & editing:** Noraini Mohamad, Nani Draman, Zainab Mat Yudin, Wan Muhamad Amir W Ahmad, Ritzzaleena Rosli Mohd Rosli, Chee Yong Chuan

